# Young adults’ views on priority health issues and their involvement in shaping responses: a qualitative exploration in South Australia

**DOI:** 10.1101/2025.07.09.25331074

**Authors:** Patience Castleton, Zahra Ali Padhani, Jaameeta Kurji, Stuart Voss, Sara Ataie-Ashtiani, Salima Meherali, Zohra S Lassi

**Author notes:** Joint first authorship (Patience Castleton, Zahra Ali Padhani). **Corresponding Author** Zohra S Lassi, Associate Professor.

## Abstract

**Background:** Adolescence is a developmental stage with a multitude of biological and social transitions as well as opportunities to set the foundation for healthy adulthood. Ensuring that young people are centred in responses to their health and wellbeing is critical for responses to be relevant, well-aligned and effective. In the context of increasing support for consumer involvement in health research, there is a need to better understand opportunities for and expectations around youth engagement.

**Methods:** Using in-depth interviews with 13 young adults (18-24 years) in South Australia, we explore what health needs are a priority, what factors are driving these issues and what needs to be done to address key concerns. We also discuss their interest in being involved in the design and delivery of interventions, their expectations and the extent to which they have had opportunities to do so. We use thematic analysis to distil key ideas and reflect on important themes across interviews.

**Results:** Mental health concerns (i.e., anxiety, depression, PTSD, eating disorders, bullying) and vaping emerged as the predominant health concerns among young adults. Lack of awareness on dangers of vaping and eating disorder, peer pressure, cost of living and financial constraints affecting ability to make healthier choices, lack of work-life balance and mental health support emerged as significant determinants influencing both options for staying healthy and accessing supports when needed. The responses emphasise effective interventions to be aligned with key determinants influencing behaviour, including early intervention, specifically in schools, to begin education and prevention efforts early. Participants also suggested free and accessible support services and programs and underscored the need for structural changes, including policy-level responses from government, to create an enabling environment for sustained impact. A key recommendation was to actively involve young adults in the design and planning of interventions to capture lived experiences. Participants shared their interest around their involvement in opportunities aligned with their interests, skills, availability and engagement preferences, but reported the existence of limited opportunities to co-design interventions at scale.

**Conclusion:** Young adults emphasised the importance of meaningful youth involvement in the design and planning of health programs. Employing the motivation and collaborative skills of young adults can help in the development of more tailored and engaging solutions aimed at promoting young adult health and well-being.

## INTRODUCTION

Adolescence is a period of rapid physical, neurological, emotional, and psychosocial transformation in an individual’s life. It presents a valuable second opportunity to consolidate the foundations of good health after the first 1000 days of life (1). Investment in young adult health and wellbeing has the potential to yield significant “triple dividends of benefits’’: improve their current health, positively shape trajectories into adulthood, and influence the well-being of future generations (2). In Australia, mental health conditions, substance use disorders and injuries are key contributors to the burden of disease in older young adults (3). Further, their active engagement in tailored health services is limited, especially amongst those living in rural communities, further decreasing their health outcomes (4, 5). Accessing affordable, culturally appropriate, peer-to-peer-led services that are informed by lived experience was identified as a major barrier to young people’s engagement with health services in the South Australian (SA) Youth Action Plan 2024-2027 (5).

Consumer-driven research was recently listed as the top priority in the *‘Australian Medical Research and Innovation Priorities 2022-2024’* (4). It acknowledges that incorporating consumers’ needs and experiences in research allows for outcomes to be better “fit-for-purpose” (4). The engagement of young adults in all aspects of activities designed for their personal and community development, including their empowerment to contribute to decisions about their personal, family, social, economic, and political development, has been recognised by the United Nations as their fundamental right (6). Whilst small efforts to increase the engagement of young adults in co-designing programs have begun in Australia, it is not well understood how and why young adults wish to be involved, or not involved, in these processes. Young respondents in the recent 2024 SA Youth Action Plan emphasised their desire to be better involved in designing and implementing the policies that affect them, expressing frustration when their views are not considered (5). However, this plan did not further investigate how to engage these young people appropriately and meaningfully in co-designing programs and services. Thus, many current efforts to engage them in co-design may fall short of meeting their unique needs and should be prioritised in research.

Meaningful engagement in co-designing programs means that individuals have the opportunity to participate in all stages of a project, including planning, action, observation, and reflection (7, 8). However, it is important to acknowledge the barriers and struggles of co-designing programs and youth engagement. For example, financial barriers with regard to reimbursements may present a significant barrier to ethically engaging young adults in programs, especially those where involvement would be lengthy and time-consuming (9). Further, concerns around the abilities of young adults to cope with the demands of involvement, lack of understanding in health programs, and boredom in sustainable contribution have been previously raised (9). Therefore, to create appropriate and engaging health programs for Australian youth and enhance youth involvement in program designing, it is important to better understand exactly how young adults want to be involved in co-designing programs

Moreover, whilst the literature is limited, the SA Youth Action Plan indicates that young South Australians are eager to engage in co-designing health programs and are invested in improving their health and wellbeing (5). Further, previous global studies have shown that young people’s engagement in health intervention designs has been enthusiastic and incredibly rewarding for both stakeholders and youth (10, 11). The outcomes of these interventions were far-reaching, and the authors claimed young people’s input was invaluable to designing interventions with lasting, positive outcomes (10, 11). It is also important that we understand the health priorities and needs of adolescents and young adults directly from them, thus allowing us to co-design interventions with their most pressing needs first. Seeking young adults’ views on what the most neglected aspects of young people’s health care are will allow us to develop a deep understanding of what interventions and programs will be of greatest benefit to them, thus ensuring the greatest benefit and engagement in co-designing programs (12, 10).

Engaging young adults in co-designing health interventions and programs is vital in ensuring the widespread adoption of these programs with far-reaching accessibility and usability. Soliciting input from direct consumers leads to increased engagement with health programs contributes to improved health outcomes for young adults, underscoring the importance of their direct involvement in research programs. Therefore, this study aimed to gain a deeper understanding of the health priorities of young adults (18 to 24 years) residing in SA and their engagement in the design of health programs.

## METHODS

To address the objective, we undertook a qualitative exploratory study adhering to the Standards for Reporting Qualitative Research checklist for reporting results (22) (**Supplementary file 1**). We conducted in-depth interviews with adolescents and young adults residing in Adelaide, South Australia. A South Australia-based youth advisory group established in March 2024 by ZSL at the University of Adelaide, consisted of five young adults, of which two provided input around key study activities, including recruitment, data collection and outputs. The duration of the study was 3 months (from October 23, 2023, until December 21, 2023).

### Setting and study participants

The study was conducted in Adelaide, South Australia and included 13 adolescents and young adults (18-24 years) identified through professional networks, organisations providing health services to young people and tertiary education institutions. Recruitment materials mainly included flyers, social media posts and personalised emails. Snowball sampling was also employed, where participants also made referrals. Those expressing interest in participating were contacted via email by the research team.

### Data collection and management

A semi-structured interview guide with prompts was followed, conducting interviews with study participants to explore what young adults felt were their most pressing health concerns and recommendations to effectively and appropriately address their needs. Interest in active roles in shaping responses through research or programmatic design was also specifically explored. Interviews were conducted in English, online or in person, depending on participant preferences. They typically lasted about 30 minutes and were conducted by trained researchers, PC and ZAP, who are female doctoral students with cultural profiles similar to those of the participants. PC is also a young adult. Interviews were audio-recorded with participant consent for transcription purposes. Notes were taken if a participant declined audio recording. Sampling continued until theoretical saturation was approached, and no substantially different ideas were identified.

### Analysis

De-identified transcripts were then analysed in dual by PC and ZAP using thematic analysis to identify patterns across cases (i.e., young adults) and to distil key ideas. These were discussed with the rest of the author team and two advisory group members (SAA and SV) who expressed an interest in the process. The decision to use thematic analysis was guided by its “theoretical flexibility”(13). A “codebook approach” was used where codes (key ideas represented in short segments of text) were used to map the analytic process following a constructivist paradigm. Rather than relying on a codebook to assess reliability and to standardise coding across coders, the codebook served as a point of discussion to chart emerging patterns during the analysis process. This approach best represents the theoretical learnings of the research team, who acknowledge that meaning is socially constructed through interaction between individuals. Analysis was performed in NVIVO software (2024/version 14) (14).

### Ethical Considerations

Ethical approval was obtained from the Human Research Ethics Committee of the University of Adelaide (H-2023-157). At the time of recruitment, participants were provided with a participant information sheet outlining what was involved in participation as well as the risks and benefits of participation to support informed decision-making. Written consent was obtained from young adults wishing to take part in the study.

## RESULTS

This study included face-to-face interviews with 13 young adults aged between 18 and 25 years old. The majority of participants were young women (n=11), and there was roughly an even split between young adults born in Australia (n=7) and those who had migrated from other countries (such as Central and South Asian and European countries). Two participants identified as members of the queer community. Most participants were enrolled in tertiary education institutions or had completed tertiary studies (n=9). Most participants still lived in their family homes with their parents or other family (n=5). All participants were working part-time or full-time in addition to being enrolled in an educational institution.

### Key health and well-being concerns and drivers

Mental health emerged as a predominant concern among participants; depression and anxiety were specifically mentioned across all interviews, while some listed eating disorders and post-traumatic stress disorder. During interviews, participants unpacked what they thought was driving these issues, often highlighting the interrelated nature of issues and the multiple levels at which determinants operated. The extracts below illustrate the dynamics between peer relationships, “lifestyle choices”, and health.

> *“There’s a whole stigma about trying to fit in and do what other people are doing, and that often includes activities such as sex and drugs, and alcohol, especially at the younger ages. Yeah, vaping now and in the younger ages like early, early teens, even earlier is crazy” (Participant # 5)*
>
> *“Bullying for young adults, like within schooling and going through schools, it happens everywhere” (Participant # 7)*

While striking an optimal balance between education or work commitments and personal life was believed to be a significant factor in maintaining mental well-being, system-level factors compounded anxiety levels. The education system, for instance, is not optimally structured to support working students. Excessive workloads and tight schedules were described as making it difficult to study and hold jobs. Additionally, schools and universities were felt to have insufficient mental health support for students.

> *“Everyone I know is overworked, over-stressed and anxious about their future and the future progression of their financial status… Schools and Universities need to plan workloads a bit better to accommodate working lifestyles as well. Just better planning and respect for the students’ timetables and workloads” (Participant # 2)*
>
> *“I think mental health is really big and being brought up within COVID… not having sound support through education… there’s not enough support. We’re burning out. We’re overworking, we can’t focus on studies, and we’re becoming stressed*.*” (Participant # 7)*

The rising trends in vaping amongst young people were also flagged as troubling. Pressure from peers, uncertainty around its impact on health and appealing and seemingly innocuous flavours of vapes were thought to be fuelling the vaping epidemic.

> *“Vaping, that’s a big thing at the moment. Lots of people, even younger than adolescents, are getting into that, and that has a big impact on them as well” (Participant #11)*.
>
> *“The reason people vape more than smoke is because you can smell when someone has smoked (cigarettes), and vaping is not like that” (Participant #9)*.

Interview participants differed in their views on responsibility, with some feeling that some young adults make “poor lifestyle choices”; others pointed out that young people are committed to staying healthy, but broader social forces, such as economic conditions and societal norms, constrain choices. As this young woman points out, social expectations and norms heavily influence behaviour.

> *“For the younger generations…we are pretty healthy and understand what it is to be healthy – [it is more] just dieting and losing weight” (Participant # 8)*.
>
> *“Australia’s kind of normalised drinking alcohol… so drinking responsibly, I think, is a bit of an issue” (Participant # 7)*.

The high cost of living was a recurring and cross-cutting theme across interviews. It was described to constrain young people’s ability to eat healthy and to stay active despite being well aware of the importance of this for good health and wellbeing. Financial strain limits opportunities for a good work-life balance and access to services, thus negatively impacting mental well-being and physical health. It evokes a sense of hopelessness as it is believed to be something out of an individual’s control. Major global crises that also had financial impacts further exacerbated existing anxieties through a disruption of support systems.

> *“The cost of things nowadays… like eating healthy*… *buying chocolate is so much cheaper than spinach, so obviously I’m going to get that. And going to the gym is so expensive*.*” (Participant # 4)*.
>
> *“A lot of the anxiety being put on us is based on stuff that you can’t really control, like finance” (Participant # 2)*.
>
> *“A big impact for young adults is their family life, which is something they can’t really control and that can hugely affect health, both physically and mentally because if your family is really poor you might not be able to afford the same food as other people, which means you don’t grow in the same way” (Participant # 12)*.

### Reflections on responses to concerns

A key recommendation that arose during discussions was to prioritise young adults in research and service design that focuses on providing mental health support for the impacts of the economic crisis they have to navigate.

> *“Everything is so different from even 10 years ago… the stakeholders and the people who are creating all these programmes, who are 50 years older than me, let alone the 60-70 years older than the young people, just don’t know. We need to be discussing it with them [young people]. What do they actually want? Don’t make decisions for them just because they’re young people, because you’re going to get such backlash, and you’re putting so much money into something that isn’t going to work*.*” (Participant # 8)*.

Ensuring that awareness campaigns were launched early was another key recommendation for programmatic responses. The nature of the recommended responses closely mirrored beliefs around root causes and key drivers of concerns. Lack of clear information on the risks associated with vaping, for instance, means that action needs to be focused on education campaigns. Emphasis was placed on including younger adolescents in these prevention efforts; as this young man suggests, educational institutions such as schools are ideal platforms for health promotion efforts.

> *“Schools [are] definitely the place for teaching young adults [about vaping]. I feel like, the earlier the better” (Participant # 4)*.

Participants varied in their views on styles of campaigns, with some suggesting fear-based approaches would be most effective.

> *“Go around to schools and talk about things that are bad for you and… because there are, especially a lot of young boys, are focusing on their health now, they wanna bulk for the gym and they wanna do this kind of thing so I guess scaring them out of vaping and putting that money towards gym,” (Participant # 8)*.

Others felt more conventional methods – websites, school-based counselling support – would be helpful. Creating supportive networks through the establishment of community groups, targeted family support and even sports-based programs were also identified as ideas for programmatic responses. Some participants were in favour of enacting bans on vapes.

Similar responses were recommended in relation to mental health concerns such as eating disorders. Information to help with recognising and managing eating disorders, with a greater in-school emphasis on developing a healthy relationship with food and exercise, was suggested.

In addition to information campaigns, responses need to address costs as a barrier to accessing support. Suggestions to leverage schools as a platform to provide free, easy-to-access mental health support were a response that particularly resonated.

> *“Mental health interventions, which I think should be introduced in schools for free and early education about things, so… educating about what should be avoided, but also what you can do, and should do, to make yourself feel better” (Participant # 12)*.
>
> *“Free counselling… cause that’s hard to come by. Like counsellors within the school” (Participant # 11)*.

Most agreed that economic burdens were not an issue that a specific program could assist with, but rather a problem that governments needed to address with specific strategies and plans. Inadequate government funding and low minimum wages were described as adding further pressure caused by the prohibitively high cost of living. The call for a “government response” reflects the view that action on structural determinants is required, as there are things that are out of young people’s control.

> *“A lot of counselling isn’t covered by Medicare and even to get a [mental] health plan is expensive… it’s not as accessible as it could be” (Participant # 4)*.

### Involvement in response formulation and expectations around engagement

The common theme from interviews was enthusiasm for more meaningful engagement in co-designing responses to key health needs. Most were motivated by the desire to contribute to efforts to keep themselves and their peers happy and healthy. The value of first-hand experience was emphasised as being critical to ensuring programmatic responses are successful in meeting young adult needs. However, hardly anyone reported being offered an opportunity to be involved in the conceptualisation or design of programmatic responses to improve young adult health.

> *I want to make a change, and I want to help people reach their best potential because I’ve been fortunate enough to have a pretty good life and pretty good upbringing, so I feel like I have a really good advantage in the world, but a lot of people are really disadvantage and don’t have that. And that’s not fair” (Participant # 7)*.
>
> *“I really see the impact that poor wellbeing and mental health has and the impact that changing it could have” (Participant # 12)*.
>
> *“I’ve struggled with some health issues in my time, so like it’s not a good feeling in yourself. It’s not great, so just making sure that everyone has a good time with themselves and knows what’s good for themselves” (Participant # 3)*.

Reflecting on expectations around involvement, young people spoke of ensuring that alignment between tasks, interests and skills. Considerations around time commitments and remuneration were also touched on.

> *“I’d like to do an education and promotion kind of programme, or like going to a talk or something, probably. I probably wouldn’t want to be part of like an education programme where I would have to educate and talk to teenagers or anything though, because I hate talking in front of people” (Participant # 6)*.
>
> *“I think I would be able to help design it and stuff. Maybe not be a whole big major part of it, but I would be more than happy to be a part of like the building block” (Participant # 9)*.
>
> *“I don’t have any skills, but yeah, if I get any task to do, I will try my best to do that” (Participant # 1)*.
>
> *“I think it depends on the vibe of it, like what they’re [the program] standing behind, what they’re wanting to achieve*.*” (Participant # 9)*.
>
> *“Unfortunately, there’s just not enough time in the day to be giving up paid hours anymore*.*” (Participant # 2)*.

Using platforms for engagement that suit young people’s preferences is also important. For some, this meant being able to engage in co-design in person rather than online.

> *“If being involved in that kind of programme meant Zoom meetings and sitting down at my desk online on my computer by myself, I would hate it; if it were an interactive in-person group meeting, then I would love it*.*” (Participant # 4)*.

## DISCUSSION

In this qualitative study, we explored the views of young adults on priority health issues and their involvement in shaping responses. Mental health concerns (i.e., anxiety, depression, PTSD, eating disorders, bullying) and vaping emerged as the predominant health concerns among young adults. Lack of awareness on the dangers of vaping and eating disorders, peer pressure around vaping, cost of living and financial constraints affecting access to services and the ability to make healthier choices, lack of work-life balance and mental health support emerged as significant determinants influencing both options for staying healthy and accessing supports when needed. The responses emphasise effective interventions to be aligned with key determinants influencing behaviour, including early intervention, specifically in schools, to begin education and prevention efforts early. Participants also suggested free and accessible support services and programs and underscored the need for structural changes, including policy-level responses from government, to create an enabling environment for sustained impact. A key recommendation was to actively involve young adults in the design and planning of interventions to capture lived experiences. Participants shared their interest around their involvement in opportunities aligned with their interests, skills, availability and engagement preferences, but reported the existence of limited opportunities to co-design interventions at scale.

This work highlighted mental health concerns (e.g. anxiety, bullying, eating disorders) and vaping as important concerns for young adults living in South Australia. According to the Australian Institute of Health and Welfare (AIHW), mental health and substance use disorders are the leading contributors to the high burden of disease among young people (15). The National Study of Mental Health and Wellbeing (NSMHWB) also reported very high levels of psychological distress among young people aged 16-24 years in the years 2020-22, with young girls (34%) experiencing more psychological distress than young boys (18%) (15).

The 2021 Household, Income and Labour Dynamics in Australia (HILDA) survey also reported high psychological distress (42.3%) and loneliness among young people aged 15-24 years (16). In addition to mental health concerns, vaping was also considered a significant issue among young people in Australia (17). Vaping was found to be the most common (49%) among young people (18-24 years) in 2022-23, which has doubled since 2019 (26%) (17). The HILDA survey also reported a higher rate of vaping among young men compared to young women in 2021 (16). According to the 2024 Youth Survey, 35% of young people voice concerns about vaping among adolescents in Australia (18).

Cost of living and financial constraints were emphasised as key determinants, playing a role in opportunities to stay healthy and access supports. According to the 2024 Youth Survey, 56% of young people identified the cost of living, and 23% identified mental health as important issues in Australia today(18). In South Australia, 54% of young people considered the cost of living a major concern in Australia(18). Data from 2023, The Advocate for Children and Young People, showed that young Australians are struggling to deal with the ongoing cost of living crisis, with 27% of study participants not seeking medical services even when unwell, 26% buying less food, and 31% skipping meals. Further, 34% of tertiary students studied had avoided purchasing supplies for their education, saving this money for other key essentials such as food and heating instead. Worryingly, 50% of young adults had also increased their level of debt to manage their financial hardships, increasing their level of stress and anxiety when required to pay this money back to friends, family and money borrowing services, such as Afterpay (19). As shown in the current study and the unwavering data, urgent action is needed to ease the financial burden among young adults of Australia, thus allowing them to live a healthy and balanced lifestyle in which they can access medical and mental health supports when needed. The Youth Survey urge the need for greater policies that tackle this crisis by increasing rates of Jobseeker services, youth allowance and rental assistance (18), with young people in the current study supporting these beliefs and further urging the need for subsidised mental health support.

Social norms at both the peer and societal levels play a role in the ability to make healthier choices. Studies suggest both social support and social norms predict health behaviours (20, 21). A study from Victoria, Australia, suggested an association between social norms with physical activity and healthy eating, likely through conformity, shared values, or social clustering (20). For example, women from underserved areas observe people around them being engaged in brisk exercise or prioritising fruits and vegetables, developing a positive attitude towards them, with an experience of social pressure to fit (20). Social norm campaigns have proven effective in multiple health promotion settings, including anti-smoking (22), traffic safety and protective measures during the COVID-19 pandemic (23).

Young adults in the current study observed that vaping and drinking behaviours are commonly started as a result of peer pressure and pressures from societal norms, therefore indicating the high influence of social norms on young adults’ behaviours. Shifting this view and developing social norm campaigns that instead highlight the prevalence of a desired behaviour, or the limited uptake of an undesired behaviour, may influence young adults to adopt similar actions. A recent meta-analysis showed that interventions aimed at reducing mental health stigma amongst young people had a significantly positive association with increased uptake for help-seeking behaviours and intentions, and additionally reduced their stigma and behaviours towards mental health (24). The current study is well supported by previous research that strongly indicates the immense ability that social norms have in changing young adults’ behaviours and perceptions and thus should be leveraged when promoting healthy choices.

Educational institutions also play a significant role in the well-being of young people; institutional policies that are responsive to the need for young people to balance education, employment and well-being are needed. Educational institutions have been seen to play significant roles in the well-being of young adults in Australia and are thus a vital setting in which health interventions should occur (25, 26). Australian studies have indicated the need for school-based mental health literacy interventions to be provided to young adults to help them find good-quality information and resources and normalise help-seeking behaviours to reduce stigma (25, 27). However, beyond this, a systematic review by McCallister et al. (28) points to the urgent need for policies around education to positively impact mental health, as the current policies typically focus on academic performance compared to mental health and well-being. This belief strongly aligns with the findings in the current study that show young adults feel increased pressures at school to achieve high academic results, whilst also failing to provide adequate mental health support (25, 27).

Extensive interest in being involved in designing responses to key health concerns was registered; however, limited opportunities were encountered. A scoping review by Sellars et al also reports on the low level of engagement of young people in health research (29). It’s important to note that the involvement of young people in the design and planning of health research is gradually growing as the field expands. Funding agencies have also placed greater emphasis on consumer representation in designing and implementing health incentives (29). However, significant barriers continue to exist at the research, organisation and funding levels, including a lack of resources, challenges in accessing young people and complex ethical approvals. These barriers often prohibit the level at which young people can engage with researchers in designing health incentives, with few opportunities provided for them despite their openness to engage. Organisations and funding bodies must, therefore, work to increase their engagement with young adults who wish to be heard and represented in health incentives.

Lived experience was emphasised as pivotal for ensuring that responses were relevant to young people and responsive to their needs. The eagerness among young people, shared in previous reports (29, 30), was promoted by their lived experience and motivation to help others. The involvement of young people provides many reciprocal benefits, such as skills development, empowerment, and social engagement, as well as increasing the feasibility, young adult friendliness, and ecological validity of the research (31, 32). Ensuring an inclusive research process requires openness to learning, strong community leadership, and trust between all research members and young people involved (33). This is an incredible opportunity for the research community to lead in this space, empowering remuneration, power sharing and leadership across sectors. Ensuring that young voices are heard across all sectors of research and policy is vital in producing initiatives that are relevant, accessible and have a great and sustainable impact.

Young people had clear expectations for engagement. These included ensuring that interests, skills and engagement platforms aligned with preferences. Considerations around their availability and ensuring appropriate remuneration for their time and contributions were also flagged as important. Despite a strong interest in health promotion and research activities, young adults in the current study acknowledged several constraints and concerns in designing and participating in health programs, including time constraints and lack of background knowledge. Young adults expressed their willingness to join a program if it were freely available, aligned with their work of interest and if familiar individuals were involved in the program. These findings align with the existing research emphasising appropriate reimbursements for time and contribution of young people invested in the decision-making initiatives (34, 35). Research additionally shows the importance of building strong, trusting relationships between young adult participants and key stakeholders to further incentivise meaningful engagement (36). Similarly to the findings in the current study, previous research shows that ensuring the interests, skills and engagement platforms align with the preferences of the young adults engaging is essential in maintaining participation (35) (36). It is, therefore, vital that young adults are provided with transparent details regarding their engagement and are appropriately reimbursed for their important insights.

### Limitations

Whilst this study provides valuable insights into the health priorities of adolescents in South Australia, it is important to note that our study has limitations. The sample included mostly females in their early 20’s who were well educated and thus may not represent the experiences and views of diverse genders and ages in SA. Further, whilst our sample was culturally diverse, the small sample size limited the diverse perspectives of many important young adults in SA, including those without tertiary education and from non-English speaking backgrounds. Additionally, most of our study came from urban settings; thus, those living in rural areas were not represented in the study outcomes. Therefore, our study sample is likely to share common concerns, and there may be additional health concerns unique to different groups not captured here.

## CONCLUSION

This study amplifies the voices of young adults in SA, revealing a pressing need to address mental health challenges, the rising prevalence of vaping, deeply influenced by social norms, lack of knowledge, peer pressure, financial constraints, and systematic barriers to care. Despite their willingness and enthusiasm to engage, young people face limited opportunities to shape the health interventions meant for them. Effective and sustainable solutions must not only align with the lives and realities of young adults but also actively involve them in the design, implementation, and evaluation of programs and policies.

We call on governments, health systems, researchers, policy makers, and educational institutions to prioritise early, school-based interventions, ensure universal access to affordable and youth-friendly mental health and substance use services, and implement structural policies that mitigate the rising cost of living and its adverse effects on health. More importantly, we urge stakeholders to create meaningful and resourced pathways for young people to co-design solutions, with appropriate remuneration, flexibility, and respect for their lived expertise. Empowering young people is not just the right thing to do; it is also key to creating health systems and communities that are fair, inclusive, and prepared for the future.

## DECLARATIONS

## Supporting information

Supplementary File 1: Checklist-Standards for Reporting Qualitative Research (SRQR)*

## Author contributions

ZSL, JK, and SM conceptualised the study. ZAP and ZSL developed the study questionnaire and received feedback from all authors. Interviews were conducted, transcribed and analysed by PC and ZAP and reviewed by ZSL. The first draft of the manuscript was written by PC and ZAP. All authors reviewed and approved the manuscript.

## Acknowledgements

None.

## Competing Interests

The authors declare no competing interests.

## Funding

This study was funded by the Robinson Research Institute Engaging Opportunity Grant 2022. ZSL (# GNT2009730) is an NHMRC Investigator Fellow.

## Data availability

All data produced in the present study are available upon reasonable request to the authors.

## Notes

### Competing Interest Statement

The authors have declared no competing interest.

### Author Declarations

The Human Research Ethics Committee of the University of Adelaide gave ethical approval for this work (H-2023-157)

